# Associations between adolescent mental health and health-related behaviours in 2005 and 2015: A population cross-cohort study

**DOI:** 10.1101/2020.03.06.20032235

**Authors:** Suzanne H. Gage, Praveetha Patalay

## Abstract

**Background:** Adolescent mental ill-health is a growing concern. There is little understanding of changes over time in the associations between mental health and health-related behaviours and outcomes such as substance use, anti-social behaviour and obesity. We investigate whether the associations between different health-related outcomes in adolescence are changing over time in two recent cohorts of adolescents born ten years apart.

**Methods:** Data from two UK birth cohort studies, the Avon Longitudinal Study of Parents and Children (ALSPAC, born 1991-92, N=5627, 50.7% female) and Millennium Cohort Study (MCS, born 2000-2, N=11318, 50.6% female) at age 14 sweeps are used. The health outcomes of focus are depressive symptom score, substance use (alcohol, smoking, cannabis and other drugs), antisocial behaviours (assault, graffiti, vandalism, shoplifting and rowdy behaviour), weight (BMI), weight perception (perceive self as overweight) and sexual activity (had sexual intercourse). Regression analyses are conducted to examine associations between these variables with cohort as a moderator to examine cohort differences.

**Results:** The directions of associations between mental-health and health-related behaviours (eg smoking) are similar over time, however, their strength across the distribution has changed. While smoking and alcohol use behaviours are decreasing in adolescents, those that endorse these behaviours in 2015 are more likely to have co-occurring mental-health than those born in 2005. Similarly, higher BMI is more strongly associated with depressive symptoms in 2015 compared to 2005.

**Conclusions:** Adverse health-related outcomes such as greater substance use, mental health difficulties and higher BMI appear to be more likely to cluster together in the more recent cohort, with implications for public health planning, service provision and lifelong disease burden.

**Implications and Contribution:** Adverse health outcomes like internalising mental ill-health, substance use and high BMI were more likely to co-occur in 2015 than 2005, albeit decreases in prevalence of substance use and antisocial behaviours and increases in internalising mental ill-health and BMI over this decade. This has implications for their causal associations and appropriate public-health action.

## Introduction

There is substantial evidence from population-based surveys and diagnostic prevalence studies that the rates of common internalising mental health difficulties such as depression and anxiety have been rising over the last few decades in adolescence (1, 2). For instance, national disorder prevalence data the UK highlights a 50% increase (3.9% to 5.8%) in emotional disorders between 2004 and 2017 (3). Findings from population based surveys in the 21^st^ Century also highlight substantial increases in internalising symptoms, for instance one study reported prevalence of 10.2% in 2009 and 13.4% in 2015 in 11-13 year olds in England (4). In parallel, there is clear evidence that the age at first smoking and drinking and prevalence of underage substance use is steadily decreasing,(5) and although evidence for antisocial behaviour is less consistent, what there is suggests decreasing trends in recent decades (3, 6, 7). For instance, in England, the Office for National Statistics reports indicate that in 1982 16% of 14 year olds described themselves as regular smokers. In 2014 this had fallen to just 4% and the observed decrease in use was particularly pronounced since the early 2000s (5).

Alongside these changes in prevalence, there is a large body of epidemiological evidence showing links between these outcomes, for example between substance use and depression (8-10), where evidence across many substances including alcohol, tobacco and cannabis consistently shows a positive association with increased rates of depression or depressive symptoms. Similarly, antisocial behaviour and substance use are known to be associated positively (i.e. they are likely to co-occur)(11) and antisocial behaviour and depressive symptoms are also more likely to co-occur in adolescence (12).

In our recent paper comparing prevalences of mental health and health-related behaviours in two cohorts of adolescents aged 14 in 2005 and 2015 (6), we found an almost two fold increase in the prevalence of high depressive symptoms and marked reductions in antisocial behaviours and lower cigarette, alcohol and drug use in 2015 compared to 2005 (6). We also observed an increase in BMI and obesity, and differences in other health related behaviours such as sleep, weight perception and sexual behaviour (6). There are a number of possible explanations for these differing changes in trends that see some outcomes increasing and others decreasing over this decade. First, these findings might indicate that the associations between these factors are dynamic, and have changed between the cohorts (13). Second, it could indicate that the associations are not causal, and that other confounding factors are responsible for any associations seen, and it is these confounding factors that have changed (14). Third, it is also possible that the associations between these outcomes may be stable over time, but changes in other explanatory factors might account for the changing trends observed.

There is limited evidence regarding changes in such associations over time, with no pre-existing research focussed on adolescence to our knowledge, which is a critical age given most of these behaviours and outcomes are first experienced in adolescence. In adults, a US study reported that the association between mental ill-health and smoking, physical inactivity and short sleep has strengthened over time (15). They hypothesise that as smoking rates have reduced in the general population, they have disproportionately reduced in those without psychological distress symptoms, meaning smoking has become over-represented among those with poor mental health. Our current investigation will aim to establish whether this is also the case for adolescent mental health and substance use behaviours.

In this paper we aim to examine a range of key adolescent outcomes including substance use (smoking, alcohol, drug-use), anti-social behaviours, internalising mental health and weight and weight perception. All of these health outcomes are often adolescent onset, and contribute greatly to disease burden and social, economic and health outcomes through the lifecourse (16-18).

As pre-registered (19), our main research question is: Have the associations between mental health and health-related behaviours changed between the two cohorts under investigation? We will investigate whether these associations in UK adolescents have changed between 2005 and 2015, using two large birth cohorts; the Avon Longitudinal Study of Parents and Children (ALSPAC, born 1991-92) and Millennium Cohort Study (MCS, born 2000-02). Our hypothesis is that associations between mental health and health-related behaviours in adolescence are dynamic and have changed between these two cohorts.

## Methods

### Participants

*Avon Longitudinal Study of Parents and Children (ALSPAC)* is a cohort born in 1991-92. ALSPAC recruited 14,541 pregnant women resident in Avon, UK with expected dates of delivery 1st April 1991 to 31st December 1992. When the oldest children were approximately 7 years of age, an attempt was made to bolster the initial sample with eligible cases who had failed to join the study originally. The total sample size for analyses using any data collected after the age of seven is therefore 15,247 pregnancies, resulting in 15,458 foetuses. Of this total sample of 15,458 foetuses, 14,775 were live births and 14,701 were alive at 1 year of age (20). The study website contains details of all the data that is available through a fully searchable data dictionary and variable search tool (http://www.bristol.ac.uk/alspac/researchers/our-data/). Ethics approval for the study was obtained from the ALSPAC Ethics and Law Committee and the Local Research Ethics Committees. Data were collected frequently via different modalities, with clinic visits and postal questionnaires having taken place in adolescence every year. This study uses data from ages 13, 14 and 15. In the current study, data were available from 6132 participants at age 14 representing 41.7% of the 14701 participants alive past 1 year. Attrition is predicted by a range of variables in ALSPAC including lower educational level, male gender, non-White ethnicity, and eligibility for free school meals.^19^

*Millennium Cohort Study (MCS)* is a cohort of 19,517 children born in 2000-02 sampled from the whole of the UK.(21) Data so far have been collected in 6 sweeps at ages 9 months, 3, 5, 7, 11 and 14 years. The <u>study website</u> (https://cls.ucl.ac.uk/cls-studies/millennium-cohort-study/) contains details regarding all the data available and information on accessing the datasets. Ethics approval for the age 14 sweep was obtained from the National Research Ethics Service Research Ethics Committee. At the age 14 sweep, 15415 families were issued into the field (those not issued due to emigration, permanent refusal, untraceability), of which 11726 families participated in the age 14 sweep (representing 60.9% of the original sample) (22). Attrition at the age 14 sweep compared to the full sample is predicted by a range of demographic variables including male gender, Black ethnicity, lower occupational and educational level and single parent family (23).

For this study, we analysed data from participants who had provided data on at least one of these outcome variables at the age 14 sweeps of the studies (depressive symptoms, smoking, alcohol, cannabis and other drugs; ALSPAC n= 6132, MCS n= 11351). Furthermore, participants without the demographic data required for increasing the comparability of the datasets (sex, ethnicity, age, maternal education and maternal age) were excluded, resulting in an analysis sample of 5627 from ALSPAC and 11318 from MCS.

There have been changes in socio-demographic characteristics of the country in the ten years between these cohorts (e.g. higher proportion ethnic minorities, higher education levels). To account for confounding due to socio-demographic differences between cohorts, we control for socio-demographic factors (sex, age, ethnicity, maternal age and parent education) in analysis.

### Measures

The measures in this study were harmonised to ensure comparability of measurement as part of a previous study (6), where we aimed to harmonise a range of mental health, health related behaviour data in these two cohorts. The harmonised measures used in this study include socio-demographic controls, mental ill-health (depressive symptoms), substance use (alcohol, smoking, cannabis and other drugs), antisocial behaviours (assault, graffiti, vandalism, shoplifting and rowdy behaviour), weight (BMI), weight perception (perceive self as overweight) and sexual activity (had sexual intercourse).

Measures that were not perfectly harmonised in this previous prevalence study (e.g. self-harm where the question relates to lifetime self-harm in ALSPAC and last 12 months in MCS), or did not derive concurrently from the same data sweep (e.g. sleep) are excluded from this study, in a deviation to this study pre-registration (19), as the differences in assessment do not make valid cross-cohort comparisons of associations possible. Details of the questions asked in each cohort and the harmonised measures used in this study are presented in supplementary Table 1.

#### Depressive symptoms

Depressive symptoms were assessed using the Short Moods and Feelings Questionnaire (24), with a cutoff score of 12 or above indicating high depressive symptoms (24).

#### Antisocial behaviour

Antisocial behaviour was assessed using questions ascertaining involvement in assault, rowdy behaviour, graffiti, vandalism and shoplifting were included in both cohorts (each coded 1 if yes), and combined to create an index of anti-social behaviours (possible range 0-5, higher score indicating engaging in higher number of antisocial behaviours). Measures are further detailed in Table S1.

#### Substance use

Variables for ever trying smoking, alcohol, cannabis and any other drugs were created, as well as variables indicating heavy drinking and weekly smoking (see Table S1 for details). To create a combined substance use index score, harmonised measures of ever trying the following substances: tobacco, alcohol, cannabis or other drugs combined to create an index score of substance use with a range of 0 (tried no substances) to 4 (used all substances). We also investigate the extent of use with tobacco (weekly smoking) and alcohol (heavy drinking).

#### Health-related behaviours

We include the harmonised variables of BMI (derived from objectively measured height and weight as measured by interviewer), weight perception (perceiving oneself as overweight) and underage age sexual activity (measure details in Table S1).

### Analysis

We first present descriptive statistics and correlations between the variables of interest in both cohorts. We examine rates of poor health behaviours/ antisocial behaviours as binary outcomes and as indices (the latter providing data on cumulative substance use/anti-social behaviours). In ALSPAC 10.4% and in MCS 1.22% of cells were missing. Multiple imputations (20 imputations) were carried out using chained equations separately in the two cohorts before combining the datasets for joint analysis. As a preliminary step in our investigation, we estimate odds of health outcomes for those with and without high depressive symptoms in each cohort.

We estimated four sets of regression models, each with depressive symptoms, substance use index score and antisocial behaviour index score as outcomes. In the first regression, we estimated the cohort interaction for each predictor separately, thus estimating the unadjusted association between each predictor and outcome and whether this is moderated by cohort. In the second regression we included a set of socio-demographic confounders, thus adjusting the association moderated by cohort accounting for these potential confounders. In the third step, we examined whether there are any non-linear associations between predictor and outcomes using a quadratic term for continuous predictors. Where non-linear associations were observed, the final model includes the non-linear term. In the fourth and final model, we mutually adjusted for all predictors (including the other outcomes) in the same model, examining whether the predictor-outcome association and the moderation by cohort are independent of the other predictors.

In order to make our comparisons between cohorts as meaningful as possible, we controlled for socio-demographic characteristics in our models to ensure differences in associations are not confounded by differences in socio-demographic characteristics between cohorts. We also use MCS sampling and attrition weights in all analyses to ensure findings from this cohort are adjusted to provide nationally representative estimates (23).

Lastly, to examine if there are any sex differences in the associations between these factors in the two cohorts we estimated three way (sex*cohort*predictor) interactions for each of our regression outcomes, and the few instances where we found some indications of sex based moderation in cohort differences, these are illustrated in figures.

## Results

We report prevalences of our main variables of interest in this investigation in Table 1. Depressive symptoms scores were higher in 2015 (mean 2015=5.72, mean 2005=4.93), mean antisocial behaviour index score was lower in 2015 (0.56, compared to 0.76 in 2005), mean substance use index score was lower in 2015 (0.59, compared to 0.66 in 2005) and mean BMI was higher in 2015 (21.58, compared to 20.32 in 2005). We report pairwise correlations between all variables of interest within each of the two cohorts in Supplementary Table S2.

**Table 1.** Descriptive statistics for mental health, antisocial behaviours, substance use and other health related behaviour outcomes in 2005 (ALSPAC) and 2015 (MCS)

| Outcome | Variable | 2005 | 2015 |
| --- | --- | --- | --- |
|  |  | ALSPAC<br>N=5627 | MCS<br>N=11318 |
|  |  | Mean or % [95% CI] | Mean or % [95% CI] |
| Depressive symptoms | Mean symptoms score | 4.93 [4.81,5.05] | 5.72 [5.57,5.86] |
|  | % above clinical cutoff | 9.0 [8.3,9.8] | 16.4 [15.5,17.3] |
| Antisocial behaviours | % Assault | 40.1 [38.5, 41.6] | 31.6 [30.4, 32.7] |
|  | % Rowdy behaviour | 11.5 [10.6,12.4] | 14.1 [13.2,14.9] |
|  | % Graffiti | 9.9 [8.9,10.9] | 2.8 [2.5,3.2] |
|  | % Vandalism | 6.2 [5.6,7.0] | 3.6 [3.1,4.1] |
|  | % Shoplifting | 8.0 [7.1,8.8] | 3.6 [3.1,4.1] |
|  | Mean antisocial behaviour index score | 0.76 [0.72,0.79] | 0.56 [0.54,0.58] |
| Substance use | Alcohol % ever drank | 52.1 [50.8,53.4] | 48.2 [47.0, 49.4] |
|  | Alcohol % heavy drinking | 3.4 [2.9,3.8] | 3.7 [3.2, 4.2] |
|  | Tobacco % ever smoked | 9.2 [8.5,10.0] | 4.7 [4.1, 5.3] |
|  | Tobacco % weekly smoking | 1.4 [1.1,1.7] | 1.8 [1.4, 2.1] |
|  | Cannabis: % tried | 4.6 [4.0,5.1] | 5.5 [4.9, 6.1] |
|  | Other drugs % tried | 0.4 [0.2,0.6] | 0.8 [0.6, 1.0] |
|  | Mean substance use index score | 0.66 [0.64,0.68] | 0.59 [0.57, 0.61] |
| Weight | Mean BMI | 20.32 [20.23,20.41] | 21.58 [21.47,21.69] |
|  | % obese | 3.8 [3.3,4.3] | 7.8 [7.1,8.5] |
| Weight Perception | % perceive overweight or very overweight | 26.4 [25.2, 27.6] | 33.6 [32.5, 34.7] |
| Sexual intercourse | % yes | 2.1 [1.8,2.5] | 2.0 [1.7,2.4] |

### Depressive symptoms as outcome

When investigating the association between rates of each different health and antisocial behaviours as predictors of depressive symptoms in both cohorts, we found that in all cases, across both cohorts, higher rates of these behaviours were associated with greater likelihood of high depressive symptoms scores, although the association between the use of an illicit drug other than cannabis and depressive symptoms was weak in 2005, with confidence intervals crossing the null. Results were broadly consistent between the two cohorts (see Figure 1) although having had sex, having assaulted someone, and having ever smoked a cigarette or drunk alcohol were more strongly associated with depressive symptoms in 2015 than in 2005.

**Fig. 1.**
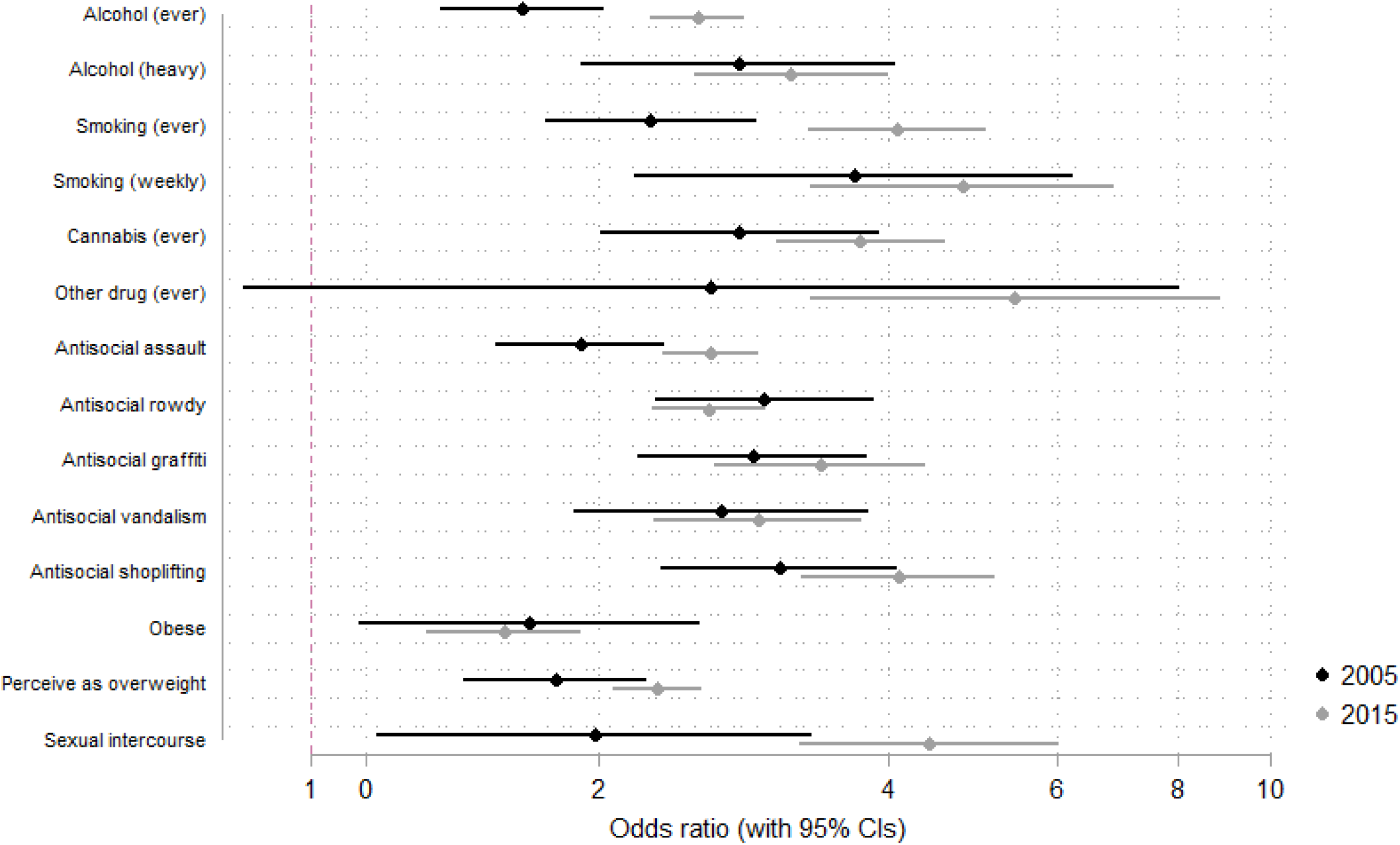
Odds of different health-related outcomes for individuals with high depressive symptoms in 2005 and 2015.

In the first regression model we observed that cohort moderates the association between all five predictors and depressive symptoms, with stronger associations at higher levels of the predictors in 2015 compared to 2005 (Figure 2). All of these remained after adjusting for confounders in step 2 (Table 2), for instance unadjusted association between antisocial behaviours and depressive symptoms coef (95% CI) = 0.92 (0.77 – 1.07), and adjusted coef (95% CI) = 1.01 (0.87-1.16). There was a non-linear association between anti-social behaviours and depressive symptoms, especially in 2015, whereby depressive symptoms were highest in the mid-range of antisocial behaviours (see Figure 2, 1A-1E).

**Table 2.**
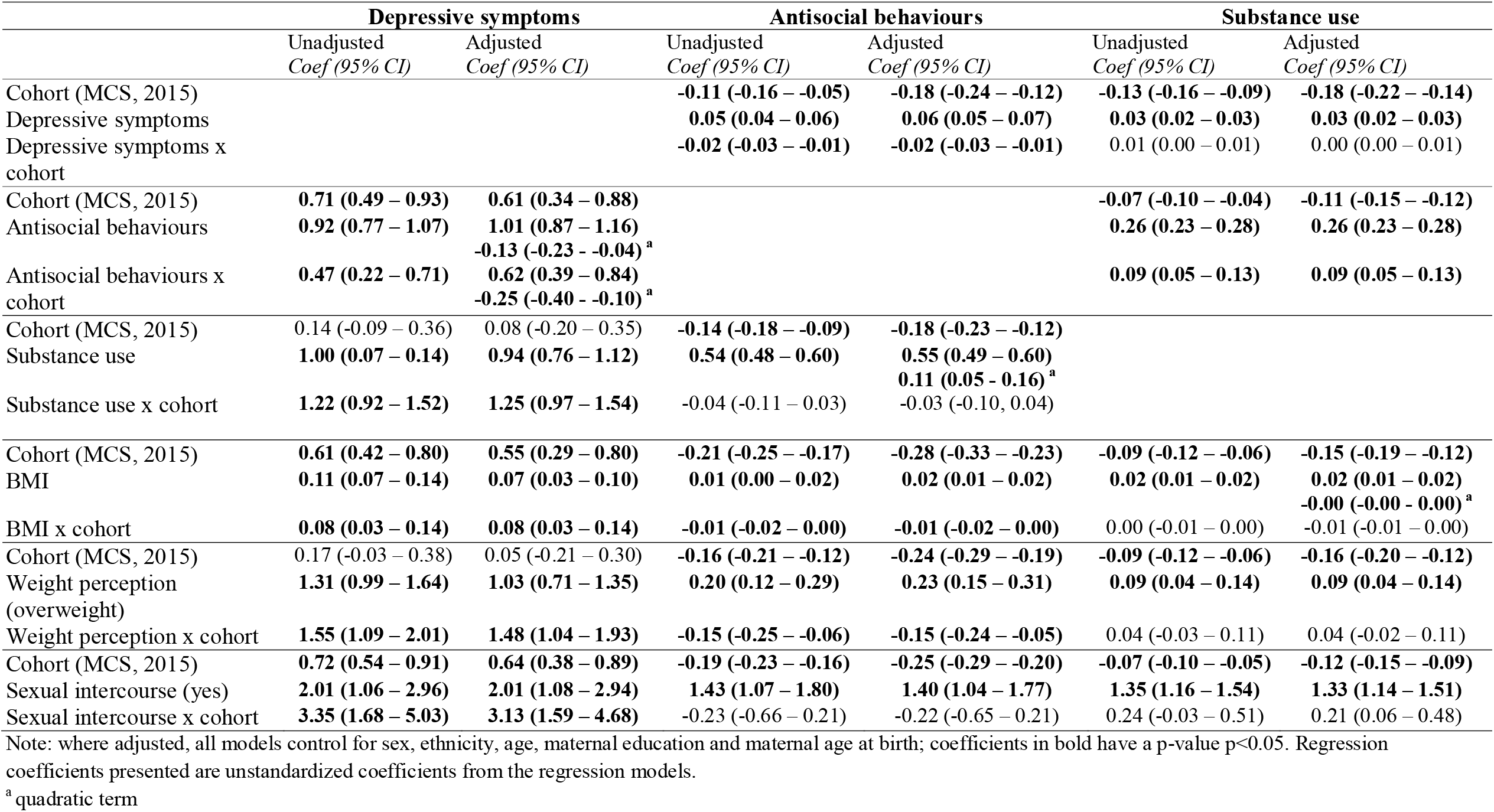
Regression coefficients from unadjusted and adjusted models for depressive symptoms, antisocial behaviours and substance use as outcomes

**Fig. 2.**
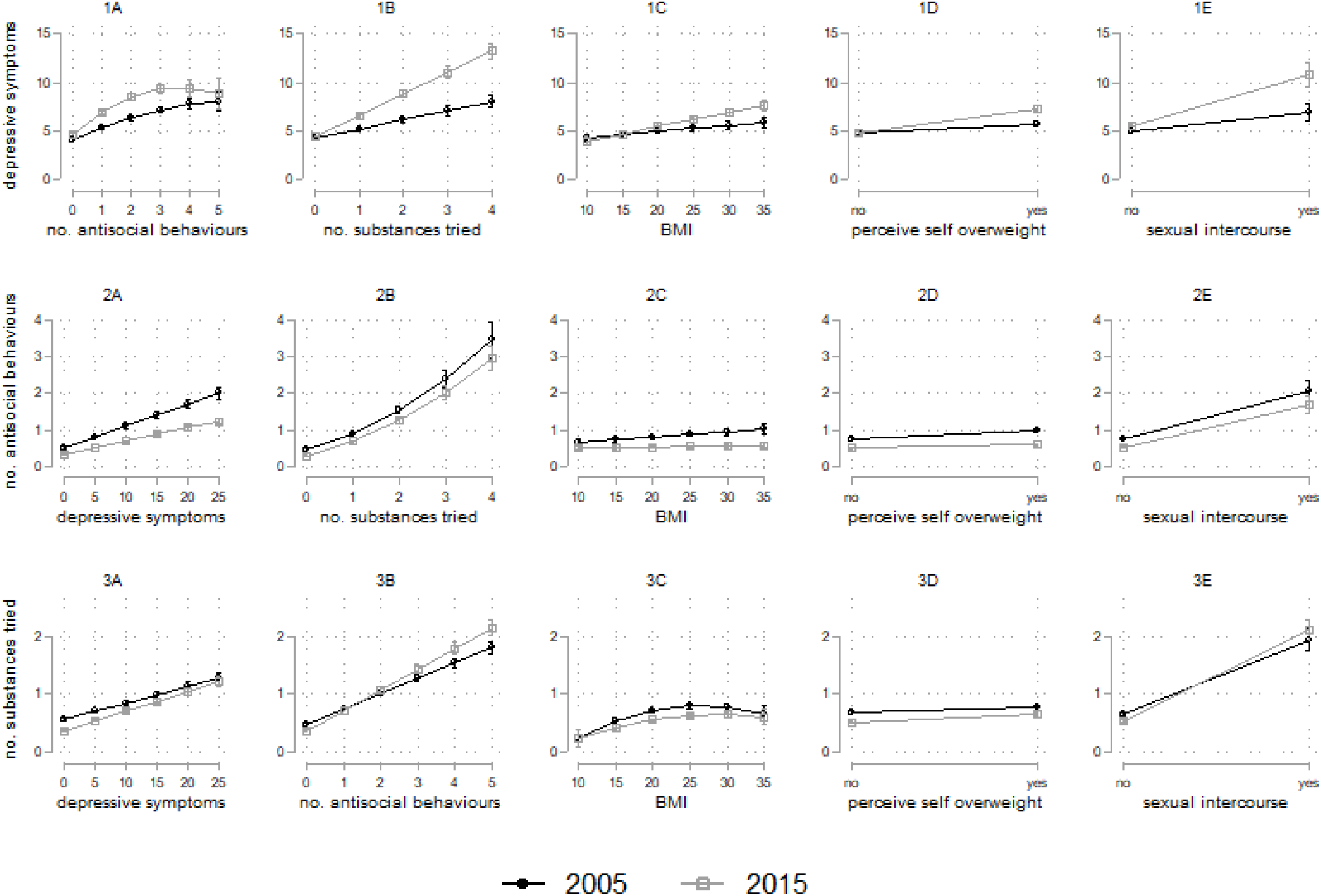
Associations between the health variables examined in 2005 and 2015. Each row of the figure corresponds to a different outcome variable (1A-E: depressive symptom score; 2A-E: number of antisocial behaviours endorsed; 3A-E number of substances tried).

In the final regression mutually adjusting for all predictors, we found evidence of a substance use by cohort interaction, where cumulative substance use was more strongly associated with depressive symptoms in 2015 than in 2005. Similarly, there was a weight perception by cohort interaction, where perceiving oneself as a higher weight was more strongly associated with depressive symptoms in 2015 than in 2005 (see supplementary Figure S1).

We investigated whether the cohort*predictor interactions were moderated by sex, and find that the cohort differences in substance use and sexual intercourse associations with depressive symptoms are primarily driven by differences in females between the cohorts, where females in 2015 demonstrate stronger associations where females with higher levels of substance use and having had sexual intercourse experience greater levels of depressive symptoms (see supplementary Fig S2a and S2b).

### Antisocial behaviour as outcome

For antisocial behaviour, in the unadjusted model we observed associations moderated by cohort for depressive symptoms, BMI and weight perception, with associations remaining similar when adjusted for confounder in Step 2 (Table 2), for example greater depressive symptoms were associated with more antisocial behaviours on average (unadjusted coef [95%CI] = 0.05 [0.04-0.06]; adjusted coef [95%CI] = 0.06 [0.05-0.07]). See Figure 2 – 2A-2E – for the associations illustrated across both cohorts.. There was a slight non-linear association between substance use and anti-social behaviour where at higher levels of both they were more strongly associated. In the final model controlling for all other predictors there was evidence of interactions between cohort and weight perception and depressive symptoms (Table 3, Figure S1). We found no evidence of moderation by sex of the cohort differences in these associations (Table S3).

**Table 3:**
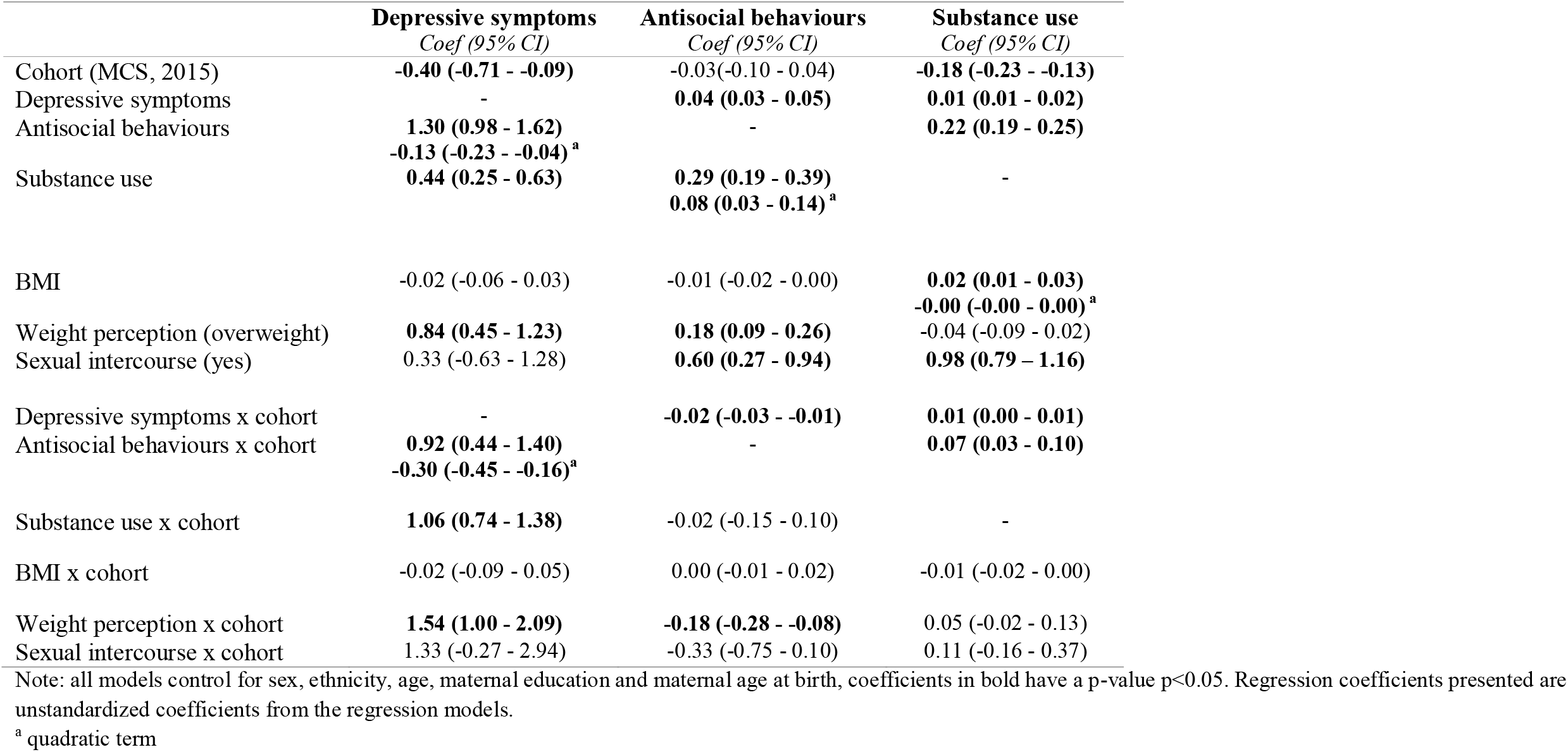
Fully adjusted regression model with all predictors and their cohort interactions included in the same model

### Substance use as outcome

For substance use, we observe associations with all the other domains considered in analysis, for instance, greater antisocial behaviours were associated with more substance use (unadjusted coef [95%CI] = 0.26 [0.23-0.28]; adjusted coef [95%CI] = 0.26 [0.23-0.28]). In the unadjusted model we observed associations moderated by cohort for depressive symptoms, antisocial behaviour and sexual intercourse and these were still observed in the adjusted model (Table 2). There was a non-linear association between BMI and substance use, where the association was stronger at above average BMI and weaker at very high BMIs (see Figure 2, 3A-3E). In our final regression, after controlling for all other variables of interest and socio-demographic factors, we found evidence of an antisocial behaviour by cohort interaction whereby at lower levels of antisocial behaviour the association was stronger in 2005 than 2015, and at higher levels of antisocial behaviour the association with substance use was stronger in 2015 than in 2005. We found no evidence of moderation by sex in the cohort differences in these associations (Table S3).

## Discussion

In this paper we examined cohort differences in the risk factor associations for several health and health related outcomes in mid-adolescence to explore whether they are changing over time or are stable between cohorts. In both cohorts, adverse outcomes in the domains investigated (i.e. greater substance use, higher antisocial behaviour, high BMI, greater depressive symptoms) were associated with higher levels of depressive symptoms, antisocial behaviours and substance use i.e. these adverse outcomes were more likely to co-occur. However, the strength of these associations varied between cohorts, with some associations also showing different patterns across their distribution. Importantly, we found that the associations between these key adolescent outcomes were often stronger among adolescents in the more recently born Millennium Cohort in 2015, compared to adolescents in 2005. This highlights that, while there may be lower prevalence of some adolescent onset health behaviours such as smoking and alcohol use in more recent cohorts, individuals with greater psychological distress and other adverse outcomes are more likely to be involved in risky health behaviours. The findings highlight the potentially changing influence of certain risk factors in more recently born cohorts, and indicate greater comorbidities or co-occuring adverse health outcomes. Understanding the dynamic nature of these associations over time is valuable for identifying causal risk factors and their changing influence as societal and environmental factors also change. This will help in identifying potential targets for interventions that can be targeted as relevant for different generations.

Turning to some of the specific findings, we found that having a higher BMI was more strongly associated with depressive symptoms in 2015 than in 2005. Part of this association was explained by the changed role of weight perception in mediating their association, with adolescents in 2015 who perceive themselves as being overweight having higher depressive symptoms than those who perceive themselves as overweight in 2005. These findings suggest that the implications of being overweight, and believing oneself to be overweight, are more severe now than they were previously, and this might be attributed to an increasing focus on appearance and perceived pressures on young people to look a certain way (25), although we cannot test this in our data. This hypothesis is supported by other work we have conducted, where we found that the proportion of adolescents are more likely to over-estimate their weight in 2015 compared to 1986 and 2005, and weight perception was more strongly associated with depressive symptoms in 2015 (26). Similarly, we observed a change between cohorts in the likelihood of antisocial behaviour and substance use in relation to depressive symptoms. For antisocial behaviour, there was a weakening in the association between depressive symptoms and the likelihood of these behaviours in 2015 compared to 2005, suggesting that the co-occurrence of these behaviours with higher psychological distress is decreasing, potentially reflecting a shift in how young people cope with difficulties, or the changing social acceptability of anti-social behaviours in adolescence. It is pertinent here that high levels of antisocial behaviours are less common in the more recent cohort. For substance use, we found lower levels of trying substances in the more recently born cohort, however, adolescents who report greater substance use, especially 3-4 substances (which are a small group, especially in 2015), have higher depressive symptoms in the recent cohort. Our general findings related to increasing co-occuring adverse outcomes are consistent with other cross-cohort studies, although these were conducted in different age groups and focussed on different outcomes. A UK-based study (which also included ALSPAC and MCS cohorts) found evidence that poor childhood mental health (at age 7) has become more strongly associated with adverse adolescent social, educational and mental health outcomes between the 1960s and 2015 (27). Similarly to a previous study of adults in the USA that found that smoking, physical inactivity and short sleep (although not heavy alcohol use) became more strongly associated with psychological distress in more recent cohorts (15), we found stronger associations between substance use and depressive symptoms in 2015 compared to in 2005. In the case of substance use the cohort difference observed in our study seemed to be primarily occurring in females, highlighting a sex specific change in vulnerability over time for this particular association.

Our study, and the above-mentioned papers, all suggest that adverse outcomes, whether substance use, mental health difficulties or health-related outcomes such as BMI appear to be more likely to cluster together in recent years compared to data from longer ago. This has consequences for disease burden throughout the rest of people’s lives, including higher likelihood of multimorbidity and widening inequalities in who experiences multiple adverse health outcomes. This may have the consequence of widening health and social disparities– people who are already experiencing some challenging circumstances may be more likely to be experiencing greater challenging circumstances than in the past. These findings have implications for clinical practice and care, for instance, the increased likelihood of co-occurring has implications for care pathways, and this evidence might be of relevance for clinicians treating current generations of adolescents with either mental health or substance use difficulties. It is also important to consider how experiencing multiple different vulnerabilities may interact in service access and treatment options. For example, there is some evidence that where individuals have comorbidities such as substance use problems and mental health problems, it is harder to get treatment for either due to the comorbidities (28).

A key strength of this investigation is the use of two large contemporary cohorts with harmonised variables available at the same age. It is important to note that one cohort, ALSPAC, is a regional cohort and MCS is a national cohort. We control for socio-demographic characteristics in our analysis and given the focus on associations between these variables, rather than prevalences, the non-nationally representative nature of one cohort is less likely to lead to bias (20). It is also important to note that missing data was higher in ALSPAC than MCS, and although we conducted multiple imputation with key socio-demographic factors and all examined variables informing the imputation to reduce bias in estimates, some estimates might remain biased due to unmeasured factors associated with missingness in each cohort and their potential association with our outcomes of interest. These analyses are cross-sectional within each cohort and it is possible that the longitudinal associations between these outcomes is also dynamic across cohorts. Understanding the early life predictors that might be changing across these cohorts will provide additional insights into the changing prevalences and associations observed between these generations.

As well as our specific research findings, we believe this paper has important implications more broadly, about the nature of how risk factors impact on outcomes. The evidence for changes in associations between cohorts suggests that risk factors are not stable over time, but rather dynamic, and a product of the societal and cultural contexts of the day. As societal and cultural factors change, these can augment or diminish the impact of certain risk factors on outcomes of interest, all of which are not independent of each other either. To assume that risk factors remain stable in terms of their ‘riskiness’ goes against the evidence. As such, where we rely on historical findings it is worth considering revisiting these in contemporary datasets, in order to ensure that any resulting public health interventions are likely to be effective. It is also important to consider that the relative impact of any one risk factor on an outcome is likely to depend on the strength of impact of all the other risk factors at play. For example, we know that the impact of genetic risk on outcomes is greater where environmental risk factors are minimised (29, 30). Similarly, broad-brush interventions that improve outcomes such as public health efforts to reduce smoking might actually increase inequality, as they are more likely to be effective for those with better resources or support (31). Using non-linear modelling to investigate patterns within associations might help to uncover examples where this is occurring and longitudinal data will help understand the longer term consequences of the increased clustering of vulnerabilities. In our data, we find evidence that gradients are steeper for the more recent cohort, suggesting that even though smoking and alcohol use behaviours are decreasing at the population level in recently born adolescents, those that are involved in these behaviours are more likely to have co-occurring mental health and other problems than in previous generations. Research to further understand the drivers of these generational trends will help understand aetiology of these important adolescent-onset health outcomes and design more effective interventions to help reduce their adverse impacts through individuals’ lives.

## Data Availability

ALSPAC and MCS cohorts are available to use upon application, and in the case of ALSPAC a fee.

http://www.bristol.ac.uk/alspac/researchers/

https://cls.ucl.ac.uk/cls-studies/millennium-cohort-study/

## Notes

### Competing Interest Statement

The authors have declared no competing interest.

### Funding Statement

This work did not receive specific funding. Praveetha Patalay is supported by a Wellcome Trust Grant (Ref: ISSF3/ H17RCO/NG1). The UK Medical Research Council and Wellcome (Grant ref: 217065/Z/19|Z) and the University of Bristol provide core support for ALSPAC. A comprehensive list of grants funding is available on the ALSPAC website (http://www.bristol.ac.uk/alspac/external/documents/grant-acknowledgements.pdf).The Millennium Cohort Study is supported by the Economic and Social Research Council and a consortium of UK government departments. The funders had no role in study design, data collection, data analysis, data interpretation, or writing of this report.

