## Supplementary file for "Associations between adolescent mental health and health-related behaviours in 2005 and 2015: A population cross-cohort study"

**This supplementary file includes:**

Table S1. Descriptions of measures in both ALSPAC and MCS and the harmonised measure used in this study.

Table S2. Correlations between all outcomes in ALSPAC (bottom left of the table) and MCS (top right of the table)

Table S3. Fully adjusted regression model with all predictors investigating sex and cohort and sex interactions

Table S4. Sensitivity analysis. Descriptives in the Millennium Cohort Study while accounting for geographical strata and clustering.

Figure S1: Associations between predictor and outcome variables, including mutual adjustment for other predictors

Figure S2a. Sex x cohort interaction for the association between number of substances used and depressive symptoms

Figure S2b. Sex x cohort interaction for the association between sexual intercourse and depressive symptoms

Table S1. Descriptions of measures in both ALSPAC and MCS and the harmonised measure used in this study.

| Outcome | ALSPAC (2005) | MCS (2015) | Harmonised variable |
|---|---|---|---|
| Depressive symptoms | [a] 13-item Short Moods and Feelings Questionnaire | [b] 13-item Short Moods and Feelings Questionnaire | Total depressive symptoms score (continuous)<br><br>Binary yes/no variable based on clinical threshold >=12<br>0 No<br>1 Yes |
| Antisocial behaviours | [b] How often in the last year have you done any of the following? (categorical)<br>- Not at all<br>- Just once<br>- 3-5 times<br>- 6+ times<br><br>Hit kicked or punched someone on purpose<br><br>Been rowdy or rude in a public place so that people complained or you got in to trouble<br><br>Written things or sprayed paint on a property that did not belong to you<br><br>Deliberately damaged or destroyed property that did not belong to you<br><br>Taken something from a shop without paying for it<br><br>Other items included skipping school, breaking and stealing from various different places, carrying a knife or weapon for protection, setting fire, stealing a vehicle, not paying correct fare on public transport, using force to steal. | [b] In the last 12 months have you: (yes/no)<br><br>Pushed or shoved /hit/slapped/punched someone?<br><br>Been noisy or rude in a public place so that people complained or got you into trouble?<br><br>Written things or spray painted on a building, fence or train or anywhere else where you shouldn't have?<br><br>On purpose damaged anything in a public place that didn't belong to you, for example by burning, smashing or breaking things like cars, bus shelters and rubbish bins?<br><br>Taken something from a shop without paying for it?<br><br>Other items included using a weapon on someone, stealing from someone, hacking into computers, sending computer viruses. | Assault<br><br>Rowdy behaviour<br><br>Graffiti<br><br>Vandalism<br><br>Shoplifting<br><br>Antisocial behaviour index score based on sum of these five harmonised binary variables. |

| | | | |
|---|---|---|---|
| Alcohol use | [a] Have you ever tried alcohol with/without your parents' permission? (yes/no)<br><br>What is the most alcoholic drinks you've had in a single evening? (continuous variable)<br><br>How many times have you done this in the last year? (continuous variable) | [b] Have you ever had an alcoholic drink? That is more than a few sips. (yes/no)<br><br>How many times have you had an alcoholic drink in the last 12 months? (7 response options from never to over 40)<br><br>Have you ever had five or more alcoholic drinks at a time? A drink is half a pint of lager, beer or cider, one alcopop, a small glass of wine, or a measure of spirits. (yes/no)<br><br>How many times have you had five or more alcoholic drinks at a time in the last 12 months?<br>- Never<br>- 1-2 times<br>- 3-5 times<br>- 6-9 times<br>- 10 or more time | Harmonised variable:<br>0 Never drank a whole drink<br>1 Nothing in last 12 months/never drank 5 or more<br>2 1-2 times drank 5 or more alcoholic drinks in one evening<br>3 3 or more times drank 5 or more alcoholic drinks in one evening<br><br>Ever drank alcohol variable (yes/no) based on recoding never drank =0 and all others =1<br><br>Heavy alcohol (yes/no) variable based on 3 or more times drank 5 or ore alcohol drinks in one evening. |
| Smoking frequency | [a] Frequency teenager has smoked cigarettes in the past 6 months:<br>- 1-3 times;<br>- >4 times;<br>- once per week;<br>- never.<br><br>Number of cigarettes smoked per week in the last 6 months for weekly users (continuous variable) | [b] Please read the following statements carefully and decide which one best describes you. Do not include electronic cigarettes (e-cigarettes).<br>- I have never smoked cigarettes;<br>- I have only ever tried smoking cigarettes once;<br>- I used to smoke sometimes but I never smoke a cigarette now;<br>- I sometimes smoke cigarettes now but I don't smoke as many as one a week;<br>- I usually smoke between one and six cigarettes a week; | Harmonised variable:<br>0 Non smoker<br>1 Occasional smoker, not weekly<br>2 Smokes 1-6 cigarettes a week<br>3 Smokes more than 6 cigarettes a week<br><br>Ever smoke variable (yes/no) based on recoding non smoker =0 and all others =1<br><br>Heavy smoker (yes/no) variable based on smoking more than 6 cigarettes a week. |

| | | | |
|---|---|---|---|
| | | - I usually smoke more than six cigarettes a week. | |
| Cannabis | [a] Have you ever used cannabis? (yes/no) | [b] Have you ever tried any of the following things?<br>Cannabis (also known as weed, marijuana, dope, hash or skunk)? (yes/no) | 0 Never used cannabis<br>1 Have tried cannabis |
| Other drugs | [a] Teenager has been offered drugs? (yes/no)<br><br><if yes to above> Teenager has used drugs other than cannabis to feel good/get high? (yes/no) | [b] Have you ever tried any of the following things?<br>Any other illegal drug (such as ecstasy, cocaine, speed)? (yes/no) | 0 Never used other drugs<br>1 Have tried other drugs |
| BMI | [a] Height and weight measured by interviewer | [a] Height and weight measured by interviewer | BMI derived from height and weight<br>Obese (0=no, 1=yes) derived using IOTF threshold |
| Weight perception | [b] How do you describe your weight?<br>- Very underweight<br>- Slightly underweight<br>- About the right weight<br>- Slightly overweight<br>- Very overweight | [b] Which of these do you think you are?<br>- Underweight<br>- About the right weight<br>- Slightly overweight<br>- Very overweight | Perceive themselves:<br>1 Underweight<br>2 About the right weight<br>3 Slightly overweight<br>4 Very overweight |
| Sexual intercourse | [b] Series of questions regarding intimate contact with someone else leading to:<br>Have you had sexual intercourse with another person in the past year? | [b] Series of questions regarding intimate contact with someone else leading to:<br>In the last 12 months have you had sexual intercourse with another young person? | 0 have not had sex<br>1 had sex |

[a] Measured via interview; [b] Measured via self completion;

Table S2. Correlations between all outcomes in ALSPAC (bottom left of the table) and MCS (top right of the table)

| **ALSPAC\MCS** | Depressive symptoms | Antisocial-rowdy | Antisocial-shoplifting | Antisocial-vandalism | Antisocial-graffiti | Antisocial-assault | Alcohol | Smoking | Cannabis | Other drugs | BMI | Weight perception | Sexual Intercourse |
|---|---|---|---|---|---|---|---|---|---|---|---|---|---|
| Depressive symptoms | - | 0.16 | 0.12 | 0.09 | 0.09 | 0.15 | 0.22 | 0.16 | 0.15 | 0.07 | 0.12 | 0.21 | 0.11 |
| Antisocial-rowdy | 0.16 | - | 0.24 | 0.31 | 0.25 | 0.26 | 0.32 | 0.18 | 0.24 | 0.13 | 0.00 | 0.02 | 0.13 |
| Antisocial-shoplifting | 0.16 | 0.34 | - | 0.26 | 0.27 | 0.16 | 0.23 | 0.22 | 0.30 | 0.16 | 0.00 | 0.03 | 0.16 |
| Antisocial-vandalism | 0.12 | 0.37 | 0.33 | - | 0.36 | 0.18 | 0.24 | 0.24 | 0.29 | 0.16 | -0.01 | 0.00 | 0.15 |
| Antisocial-graffiti | 0.16 | 0.35 | 0.33 | 0.39 | - | 0.15 | 0.20 | 0.22 | 0.24 | 0.13 | 0.00 | 0.02 | 0.11 |
| Antisocial-assault | 0.14 | 0.25 | 0.22 | 0.26 | 0.25 | - | 0.22 | 0.11 | 0.16 | 0.06 | 0.00 | 0.00 | 0.09 |
| Alcohol use | 0.15 | 0.28 | 0.24 | 0.21 | 0.24 | 0.20 | - | 0.31 | 0.37 | 0.17 | 0.07 | 0.08 | 0.24 |
| Smoking | 0.15 | 0.27 | 0.27 | 0.19 | 0.24 | 0.16 | 0.37 | - | 0.49 | 0.27 | 0.03 | 0.05 | 0.30 |
| Cannabis | 0.11 | 0.19 | 0.27 | 0.19 | 0.20 | 0.14 | 0.34 | 0.44 | - | 0.33 | 0.03 | 0.03 | 0.32 |
| Other drugs | 0.02 | 0.03 | 0.08 | 0.02 | 0.04 | 0.02 | 0.13 | 0.23 | 0.26 | - | 0.00 | 0.01 | 0.18 |
| BMI | 0.08 | 0.02 | 0.03 | 0.03 | 0.01 | 0.04 | 0.09 | 0.05 | 0.03 | 0.01 | - | 0.65 | 0.02 |
| Weight Perception | 0.11 | 0.04 | 0.05 | 0.03 | 0.05 | 0.06 | 0.08 | 0.07 | 0.02 | 0.01 | 0.63 | - | 0.02 |
| Sexual intercourse | 0.06 | 0.14 | 0.17 | 0.14 | 0.12 | 0.10 | 0.23 | 0.32 | 0.26 | 0.13 | 0.04 | 0.02 | - |

Table S3. Fully adjusted regression model with all predictors investigating sex and cohort and sex interactions

| | **Depressive symptoms** *Coef (95% CI)* | **Antisocial behaviours** *Coef (95% CI)* | **Substance use** *Coef (95% CI)* |
|---|---|---|---|
| Cohort (MCS, 2015) | -0.56 (-0.96 - -0.15) | -0.06 (-0.16 – 0.03) | -0.14 (-0.20 - -0.08) |
| Sex (female) | 0.94 (0.55 – 1.33) | -0.20 (-0.30 - -0.11) | 0.00 (-0.07 – 0.06) |
| Depressive symptoms | - | 0.05 (0.03 – 0.06) | 0.01 (0.00 – 0.02) |
| Antisocial behaviours | 0.77 (0.36 – 1.18) | - | 0.22 (0.18 – 0.25) |
| Substance use | 0.31 (0.07 – 0.55) | 0.47 (0.39 – 0.55) | - |
| BMI | -0.02 (-0.09 – 0.04) | -0.02 (-0.03 – 0.00) | 0.02 (0.01 - 003) |
| Weight perception (overweight) | 0.56 (0.02 – 1.11) | 0.15 (0.01 – 0.30) | -0.09 (-0.18 – 0.01) |
| Sexual intercourse (yes) | 0.50 (-0.74 – 1.73) | 0.70 (0.20 – 1.19) | 0.97 (0.69 – 1.24) |
| Sex x cohort | 0.04 (-0.16 – 0.96) | 0.03 (-0.08 – 0.15) | -0.06 (-0.14 – 0.02) |
| Depressive symptoms x cohort | - | -0.01 (-0.03 – 0.00) | 0.00 (-0.01 – 0.01) |
| Antisocial behaviours x cohort | 1.04 (0.44 – 1.64) | - | 0.08 (0.03 – 0.13) |
| Substance use x cohort | 0.29 (-0.10 – 0.69) | 0.06 (-0.04 – 0.16) | - |
| BMI x cohort | 0.04 (-0.05 – 0.13) | 0.01 (-0.01 – 0.03) | -0.01 (-0.02 – 0.00) |
| Weight perception x cohort | 0.61 (-0.13 – 1.36) | -0.19 (-0.35 - -0.02) | 0.08 (-0.03 – 0.19) |
| Sexual intercourse x cohort | -0.63 (-2.42 – 1.17) | -0.06 (-0.67 – 0.54) | 0.08 (-0.34 – 0.50) |
| Depressive symptoms x sex | - | -0.01 (-0.02 – 0.01) | 0.00 (-0.01 – 0.01) |
| Antisocial behaviours x sex | 0.79 (0.13 – 1.44) | - | 0.01 (-0.03 – 0.06) |
| Substance use x sex | 0.34 (-0.03 – 0.71) | -0.01 (-0.11 – 0.09) | - |
| BMI x sex | 0.04 (-0.05 – 0.13) | 0.01 (-0.01 - 003) | -0.01 (-0.03 – 0.00) |
| Weight perception x sex | 0.58 (-0.14 – 1.29) | 0.04 (-0.13 – 0.22) | 0.08 (-0.03 0.20) |
| Sexual intercourse x sex | -0.43 (-2.29 – 1.44) | 0.01 (-0.65 – 0.66) | 0.03 (-0.33 – 0.38) |
| Depressive symptoms x cohort x sex | - | -0.01 (-0.02 – 0.01) | 0.01 (0.00 – 0.02) |
| Antisocial behaviours x cohort x sex | 0.33 (-0.67 – 1.34) | - | -0.03 (-0.10 – 0.05) |
| Substance use x cohort x sex | 1.26 (0.66 – 1.87) | -0.17 (-0.30 - -0.03) | - |
| BMI x cohort x sex | -0.11 (-0.24 – 0.01) | -0.01 (-0.04 – 0.01) | 0.01 (-0.01 – 0.02) |
| Weight perception x cohort x sex | 1.27 (0.25 – 2.30) | 0.03 (-0.17 – 0.19) | -0.04 (-0.19 – 0.11) |
| Sexual intercourse x cohort x sex | 3.39 (0.80 – 6.59) | -0.47 (-1.27 – 0.34) | 0.04 (-0.49 – 0.56) |

Table S4. Sensitivity analysis. Descriptives in the Millennium Cohort Study while accounting for geographical strata and clustering.

| **Outcome** | **Variable** | **MCS (accounting for stratified and clustered sampling design- sensitivity check)** | **MCS (weighted but not accounting for strata and clusters-main analysis)** |
|---|---|---|---|
| | | Mean or % [95% CI] | Mean or % [95% CI] |
| Depressive symptoms | Mean symptoms score | 5.72 [5.55, 5.89] | 5.72 [5.57,5.86] |
| | % above clinical cutoff | 16.2 [15.2, 17.1] | 16.4 [15.5,17.3] |
| Antisocial behaviours | % Assault | 31.6 [30.4, 32.7] | 31.6 [30.4, 32.7] |
| | % Rowdy behaviour | 14.1 [13.2,14.9] | 14.1 [13.2,14.9] |
| | % Graffiti | 2.8 [2.5,3.2] | 2.8 [2.5,3.2] |
| | % Vandalism | 3.6 [3.1,4.1] | 3.6 [3.1,4.1] |
| | % Shoplifting | 3.6 [3.1,4.1] | 3.6 [3.1,4.1] |
| | Mean antisocial behaviour index score | 0.56 [0.53,0.58] | 0.56 [0.54,0.58] |
| Substance use | Alcohol % ever drank | 48.2 [46.3, 50.0] | 48.2 [47.0, 49.4] |
| | Alcohol % heavy drinking | 3.7 [3.2, 4.2] | 3.7 [3.2, 4.2] |
| | Tobacco % ever smoked | 4.7 [4.1,5.4] | 4.7 [4.1, 5.3] |
| | Tobacco % weekly smoking | 1.8 [1.4, 2.1] | 1.8 [1.4, 2.1] |
| | Cannabis:% ever tried | 5.5 [4.9, 6.1] | 5.5 [4.9, 6.1] |
| | Other drugs % ever tried | 0.8 [0.6, 1.0] | 0.8 [0.6, 1.0] |
| | Mean substance use index score | 0.59 [0.57, 0.62] | 0.59 [0.57, 0.61] |
| Weight | Mean BMI | 21.58 [21.48, 21.69] | 21.58 [21.47,21.69] |
| | % obese | 7.8 [7.1, 8.5] | 7.8 [7.1,8.5] |
| Weight Perception | % perceive overweight or very overweight | 33.6 [32.5, 34.6] | 33.6 [32.5, 34.7] |
| Sexual intercourse | % yes | 2.0 [1.6,2.4] | 2.0 [1.7,2.4] |

Figure S1: Associations between predictor and outcome variables, including mutual adjustment for other predictors in 2005 and 2015. Each row of the figure corresponds to a different outcome variable (1A-E: depressive symptom score; 2A-E: number of antisocial behaviours endorsed; 3A-E number of substances tried).

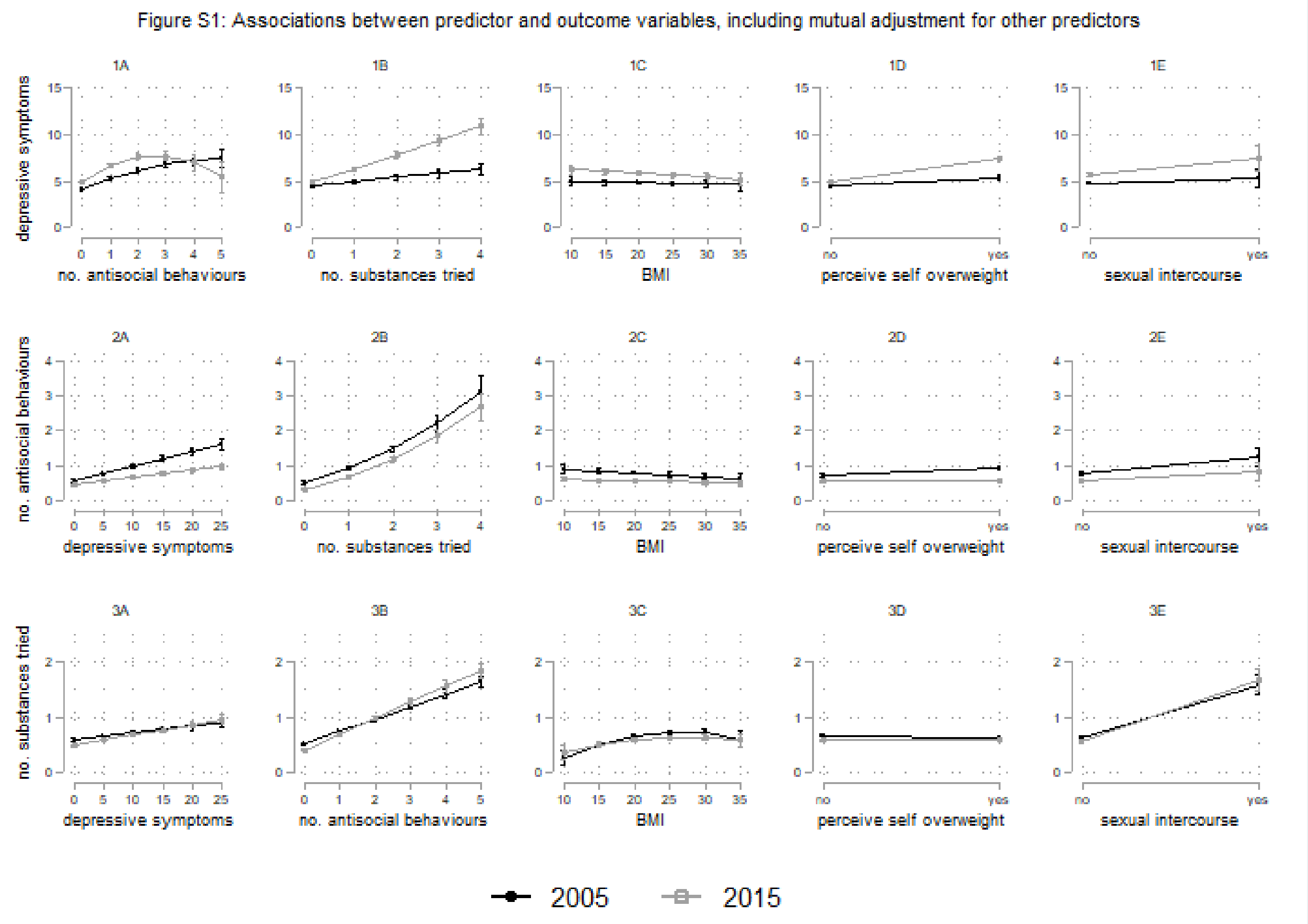

Figure S2a: Sex x cohort interaction for the association between number of substances used and depressive symptoms

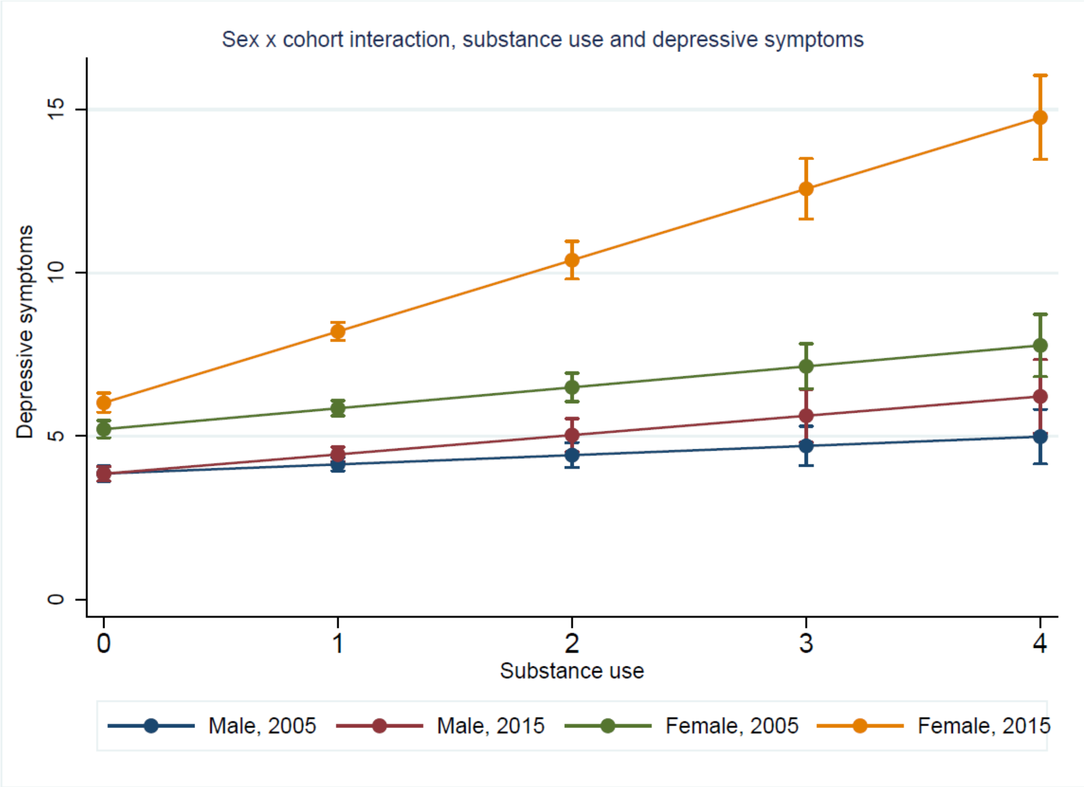

Figure S2b: Sex x cohort interaction for the association between sexual intercourse and depressive symptoms

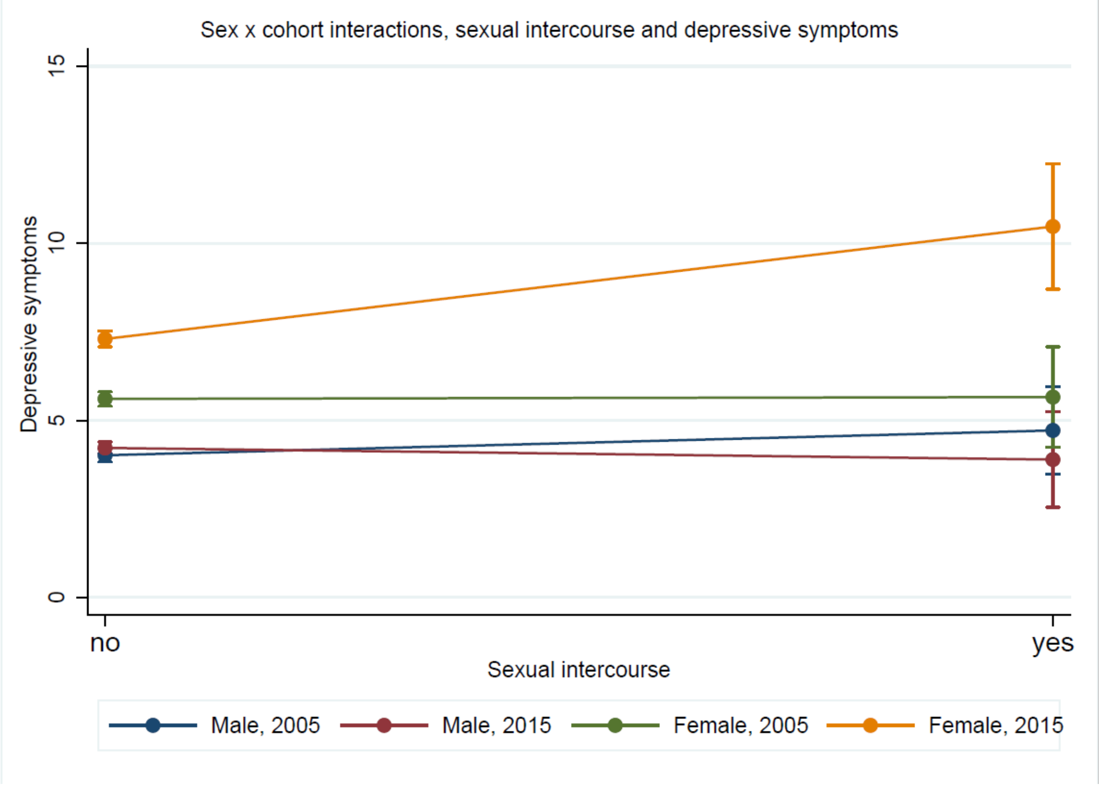
